# Demographic Composition of Participants in Sex Chromosome Aneuploidy Studies across the Globe: a 20-year Systematic Review

**DOI:** 10.1101/2025.06.17.25329799

**Authors:** Karli S Swenson, Samantha Bothwell, Anastasia Zhivotov, Amanda Sieverts, Shalika Devireddy, Kira Shuff, Kayla Nocon, Alexandra Carl, Kayla Molison, Lidia Grzybacz, Brisa Avila, Chijioke Ikomi, Lilian Cohen, Ellie Svoboda, Shanlee Davis

## Abstract

**Background:** Sex chromosome aneuploidies (SCAs), including Klinefelter syndrome (47,XXY), Turner syndrome (45,X), XYY syndrome, trisomy X (47,XXX), and rarer tetrasomies and pentasomies, affect approximately 1 in 400 births and are associated with a wide range of developmental, cognitive, and physical health outcomes. While clinical research on SCAs has expanded over the past two decades, it is unclear whether the populations included in these studies reflect the demographic diversity of those affected. Assessing representation is critical to ensuring research findings are generalizable and applicable to diverse patient populations.

**Methods:** We conducted a systematic review of global clinical research on SCAs published in English between January 2004 and May 2024. Searches were performed in Ovid MEDLINE® ALL, Embase, and Web of Science. Studies were included if they enrolled ≥10 participants and excluded if they were case reports, reviews, or meta-analyses. We extracted data from 1,474 studies on geographic location, participant karyotypes, and demographic metrics, including race, ethnicity, and socioeconomic status (SES) reported. Trends in demographic reporting were examined over time and by geographic region. For U.S.-based studies reporting race/ethnicity, we compared pooled participant demographics to national census data.

**Results:** SCA research is concentrated within a small number of geographic areas, primarily in Europe (51.4%) and the U.S. (23.6%). Reporting rates of race or ethnicity for U.S. papers increased over the 20-year observation period, with an average increase of 1.5% ± 0.4% per year (p = 0.003), peaking in 2024 with 61.4% of U.S.-based papers presenting demographics. When reported, studies consistently overrepresented non-Hispanic White (p<0.001) and college-educated (p<0.001) participants relative to U.S. census benchmarks.

**Conclusions:** This systematic review reveals persistent gaps in the demographic reporting and representation of participants in SCA research. Even in the U.S., where population diversity is high, published studies do not reflect the expected racial, ethnic, and socioeconomic makeup of affected individuals. To ensure that research findings are equitable and clinically relevant, future studies should adopt standardized demographic reporting and prioritize inclusive enrollment strategies to reflect the full spectrum of individuals with SCAs.

## INTRODUCTION

Sex chromosome aneuploidies (SCAs) are a group of genetic conditions characterized by the presence of an extra or a missing sex chromosome. Together, SCAs affect approximately 1 in 400 live births and include Klinefelter syndrome (47,XXY), Turner syndrome (TS, 45,X), trisomy X (47,XXX), XYY syndrome, 48,XXYY, and other rarer tetrasomies and pentasomies.^1,2^ These conditions affect individuals of any sex and are associated with a range of neurodevelopmental, cognitive, reproductive, and physical health outcomes.^3,4^ While the volume of research on SCAs has grown, it remains unclear whether the participants included in clinical studies are representative of the broader population affected by these conditions.

Representation in clinical research is critical to ensuring that findings are generalizable, clinical guidance is evidence-based for diverse populations, and existing health disparities are not inadvertently reinforced. Race, ethnicity, and socioeconomic status (SES) have been shown to influence access to care, participation in research, and health outcomes across many conditions.^5^ The challenges of ensuring representative research samples are magnified in rare disease research, where small study populations limit opportunities to achieve demographic diversity. While some conditions may be enriched in certain populations due to founder effects or environmental exposures, SCAs are believed to occur with equal frequency across all populations.^6^ However, SCAs are significantly underdiagnosed: an estimated 75% of individuals with 47,XXY and up to 90% with 47,XXX or 47,XYY remain undiagnosed.^7^ Importantly, disparities in access to genetic testing have been well documented, with studies showing that individuals from minoritized racial and ethnic groups are less likely to be offered or accept genetic screening.^8^ This creates the potential for bias in who is diagnosed and who ultimately participates in research. However, the increasing uptake of prenatal cell-free DNA (cfDNA) provides an opportunity to shift the landscape, enabling earlier and more equitable diagnosis, if care pathways are optimized.

The objective of this systematic review is to evaluate the demographic composition of participants included in original clinical research studies on SCAs. Specifically, we assess reporting of race, ethnicity, SES, and geographic location in peer-reviewed publications over the past two decades. By characterizing demographic trends over time and comparing U.S. study populations to national census benchmarks, we aim to determine whether the current literature accurately reflects the populations affected by SCAs. These results have implications for clinical care and future research priorities.

## METHODS

This systematic review was conducted in accordance with PRISMA guidelines, and the search strategy was registered with Prospero (ID# CRD42024550312) prior to initiating the literature search.^9^ A comprehensive search strategy including both controlled vocabulary and free-text terms related to SCAs was developed in collaboration with a health sciences librarian (ES) and peer reviewed by a second librarian (see Supplemental Table 1). The search was performed on May 21, 2024, across the following databases: Ovid MEDLINE® ALL (1946–May 20, 2024), Embase (Embase.com, 1974–May 21, 2024), and Web of Science. Results were limited to human studies published in English between January 2004 and May 2024. Case reports, reviews, meta-analyses, editorials, commentaries, letters, news, and meeting abstracts were excluded. The term ‘parsonage’ was excluded to remove irrelevant records related to Parsonage-Turner Syndrome.

The search retrieved 19,031 citations, which were downloaded into EndNote version 21 and uploaded into Covidence, a systematic review management platform.^10^ A total of 8,451 duplicates were identified and removed by EndNote or Covidence. The remaining 10,580 citations were screened with titles and abstracts. 8,651 were removed for not meeting inclusion criteria (i.e. human studies on SCAs with at least 10 participants), leaving 1,929 for full text review (Figure 1).

**Figure 1.**
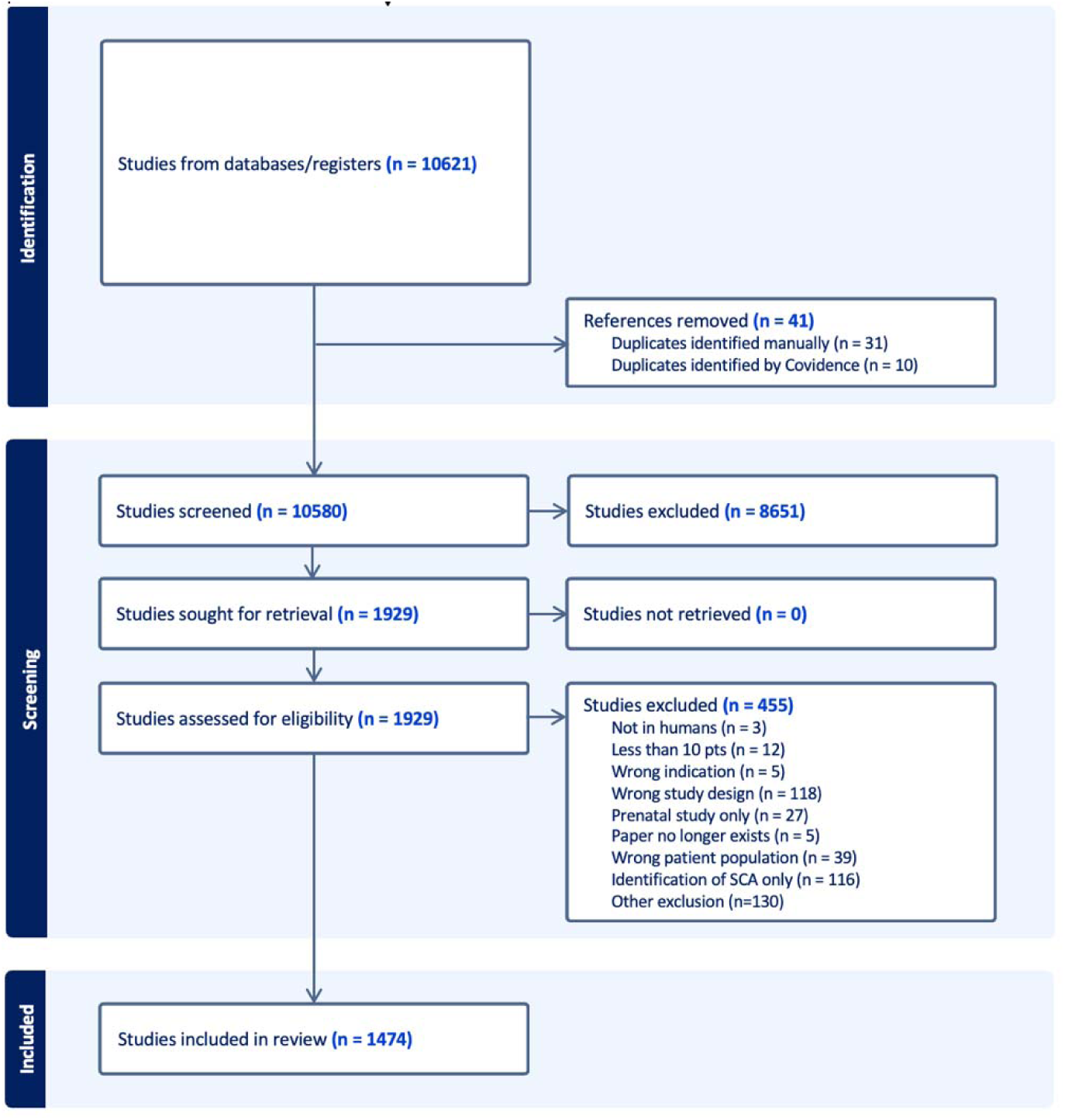
PRISMA flow diagram of included studies.

### Article review and data extraction

Title and abstract screening were performed independently by two reviewers (KSS, AZ, AS, SD, KS, AEC, LG, KN, KM). Discrepancies were resolved by consensus or adjudicated by KSS. Of the 1,929 articles identified for full-text review, a reviewer (AZ, AS, KS) retrieved the full-text manuscripts via open access or InterLibrary Loan. Full texts were independently reviewed by two reviewers to determine inclusion eligibility, with disagreements resolved by KSS. 468 manuscripts were excluded at the full text review stage for not meeting inclusion criteria.

The final dataset comprised 1,474 manuscripts. Data were extracted using an author-developed template that included publication characteristics (title, year), author information (location, contact), geographic region based on location of the corresponding author, study design, SCA conditions included, general topic studied, number of participants, and participant race, ethnicity, and any socioeconomic metrics (e.g. education, income, insurance status, Hollingshead index). We used a broad inclusion methodology, considering any mention of culturally relevant race or ethnicity indicators (e.g., "all Nigerian" would be counted as reporting race/ethnicity). Each manuscript extraction was completed by one reviewer (KSS, AZ, AS, SD, KS, AEC, LG, KN, KM), with the first 20 extractions for each reviewer verified by KSS. Any questions or discrepancies were resolved through discussion with KSS.

### Statistical Analysis

Descriptive statistics are presented as medians with interquartile ranges (IQR) for continuous variables and as n (%) for categorical variables. Geographic distributions of included studies are visualized globally and by U.S. states using the usmap R package, version 0.7.1. The proportion of studies reporting race, ethnicity, and/or any SES measure were calculated for all and then stratified by specific SCA groups as well as U.S./International non-U.S. studies to identify global hubs for SCA research. To contextualize geographic diversity in our analysis, we grouped countries based on both geographic proximity and shared socioeconomic or healthcare system characteristics as determined by the author group. For example, Northern European countries such as Sweden, Denmark, and Norway were grouped together given their similar public health infrastructures, population demographics, and research practices. This approach allowed for a more meaningful interpretation of demographic trends and study generalizability across regions with comparable societal structures. Temporal trends in race/ethnicity and SES reporting are visualized from 2004 to 2024. We evaluated linear regressions to assess if an increase in reporting occurred over time as well as general publication rates for SCA research. In 2013, NIH reporting guidelines mandated race and ethnicity reports be included in publication of clinical research.^13^ To assess if there was a difference after the guidelines were released, race/ethnicity reporting rates prior to 2013 are compared to after to 2013.

For studies within the U.S. reporting race and/or ethnicity, we conducted a meta-analysis of proportions to estimate the pooled representation of racial and ethnic groups across included studies. Specifically, for papers between 2014 and 2024, the weighted average of the percent of SCA participants who were non-Hispanic White, Hispanic, Black or African American, and Asian participants. These racial/ethnic groups were chosen based on frequency of reporting across studies. Weighted averages were calculated separately for each racial/ethnic category as the common effect across studies using the metaprop function in the meta R package, version 7.0-0.^11^ The inverse variance method was applied to weight the studies proportional to their sample size. I^2^ measures are reported as a measure of heterogeneity across studies. Weighted proportions are compared to the U.S. Census estimates using one-sample z-tests.^12^ We conducted a similar analysis for education level for the proportion of parents with a college degree or greater. A type 1 error rate of 0.05 was assumed for all analyses. Analyses were performed in R, version 4.4.1.

## RESULTS

### Study Characteristics

1,474 manuscripts are included in the analysis. Most papers included Turner syndrome (45,X/variants) (68.2%), representing a total of 163,816 participants (Table 1). Papers on 47,XXY were the next most frequent (29.4%), while all other SCAs were each represented in 2.0% - 5.6% of studies. Studies included a median of 55 participants (IQR 26-108). Almost all papers reported on observational studies. Overall, cardiac/cardiometabolic topics were the most common (15.9%), followed by neurodevelopment/psych/mental health (15.7%), and physical phenotype (14.1%), although there was variability by karyotype (Supplemental Table 2). The number of SCA papers published annually increased during the study period, averaging 4.7±0.5 more papers published year over year (p < 0.001, Figure 1).

**Table 1.** Description of included studies.

|  | Number of papers |
| --- | --- |
| **Karyotype** □ | Overall (n=1,474) |
| Turner syndrome | 1006 (68.2%) |
| 47,XXY | 434 (29.4%) |
| 47,XYY | 83 (5.6%) |
| 47,XXX | 81 (5.5%) |
| 48,XXYY | 29 (2.0%) |
| Other Tetra and Pentasomies | 36 (2.4%) |
| **Median n of participants per study** | 55 [26, 108] |
| Turner syndrome | 57 [28, 113] |
| 47,XXY | 44 [23, 85] |
| 47,XYY | 24 [13.5, 45] |
| 47,XXX | 27 [15, 35] |
| 48,XXYY | 19 [10, 25] |
| Other Tetra and Pentasomies | 17 [6, 36] |
| **Study Design** |  |
| Cross sectional study | 550 (37.3%) |
| Cohort study | 313 (21.2%) |
| Chart review | 231 (15.7%) |
| Case control study | 216 (14.7%) |
| Randomized controlled trial | 45 (3.1%) |
| Non-randomized experimental study | 40 (2.7%) |
| Case series | 23 (1.6%) |
| Basic science | 22 (1.5%) |
| Qualitative research | 22 (1.5%) |
| Other | 12 (1.2%) |
| **Topic of study** |  |
| Cardiac/cardiometabolic | 235 (15.9%) |
| Neurodevelopment/psych/mental health | 232 (15.7%) |
| Physical phenotype | 208 (14.1%) |
| Other or multiple | 194 (13.2%) |
| Growth treatment | 149 (10.2%) |
| Fertility/fertility treatment | 145 (9.9%) |
| Genetics/epigenetics | 112 (7.7%) |
| Sex steroid hormones/hormone treatment | 101 (6.9%) |
| Quality of life/wellbeing | 82 (5.6%) |
| Basic science | 16 (1.1%) |
| **Publication year** |  |
| Prior to 2004 | 1 (0.1%) |
| 2004 - 2008 | 261 (17.7%) |
| 2009 - 2013 | 324 (22.0%) |
| 2014 - 2018 | 348 (23.6%) |
| 2019 - 2023 | 509 (34.5%) |
| 2024 | 31 (2.1%) |
| **Age of participants** □ |  |
| Newborns (<1) | 172 (11.7%) |
| Toddlers (1-2) | 275 (18.7%) |
| Children (3-11) | 771 (52.3%) |
| Adolescents (12-17) | 896 (60.8%) |
| Young adults (18-29) | 944 (64.0%) |
| Middle adults (30-49) | 701 (47.6%) |
| Older adults (50-74) | 271 (18.4%) |
| Elderly (75+) | 64 (4.3%) |
| **Funding source** □ |  |
| No funder listed | 645 (43.8%) |
| Public funding | 522 (35.4%) |
| Institutional funding | 213 (14.5%) |
| Nonprofit funding | 226 (14.4%) |
| Industry funding | 130 (8.8%) |
| Other | 16 (1.0%) |

Data are presented as N (%) or median (IQR).
□ Categories are not mutually exclusive, therefore percentages do not add up to 100%.

### Global Distribution of SCA Research

Visualizing the number of papers by global region revealed most research being conducted in Europ (51.4%) followed by the U.S. (23.6%) (Figure 2). There are almost no publications from Sub-Saharan Africa, and fewer than 22% were conducted in Latin American, Asian, or African countries. In 47,XXX, 47,XYY, tetrasomy, and pentasomy conditions, 85.7% of the studies were conducted in the U.S. or Western Europe. While 8.9% and 5.8% of all TS papers were from Latin America and Japan respectively, these regions had no papers including 47,XYY and only two including 47,XXX.

**Figure 2.**
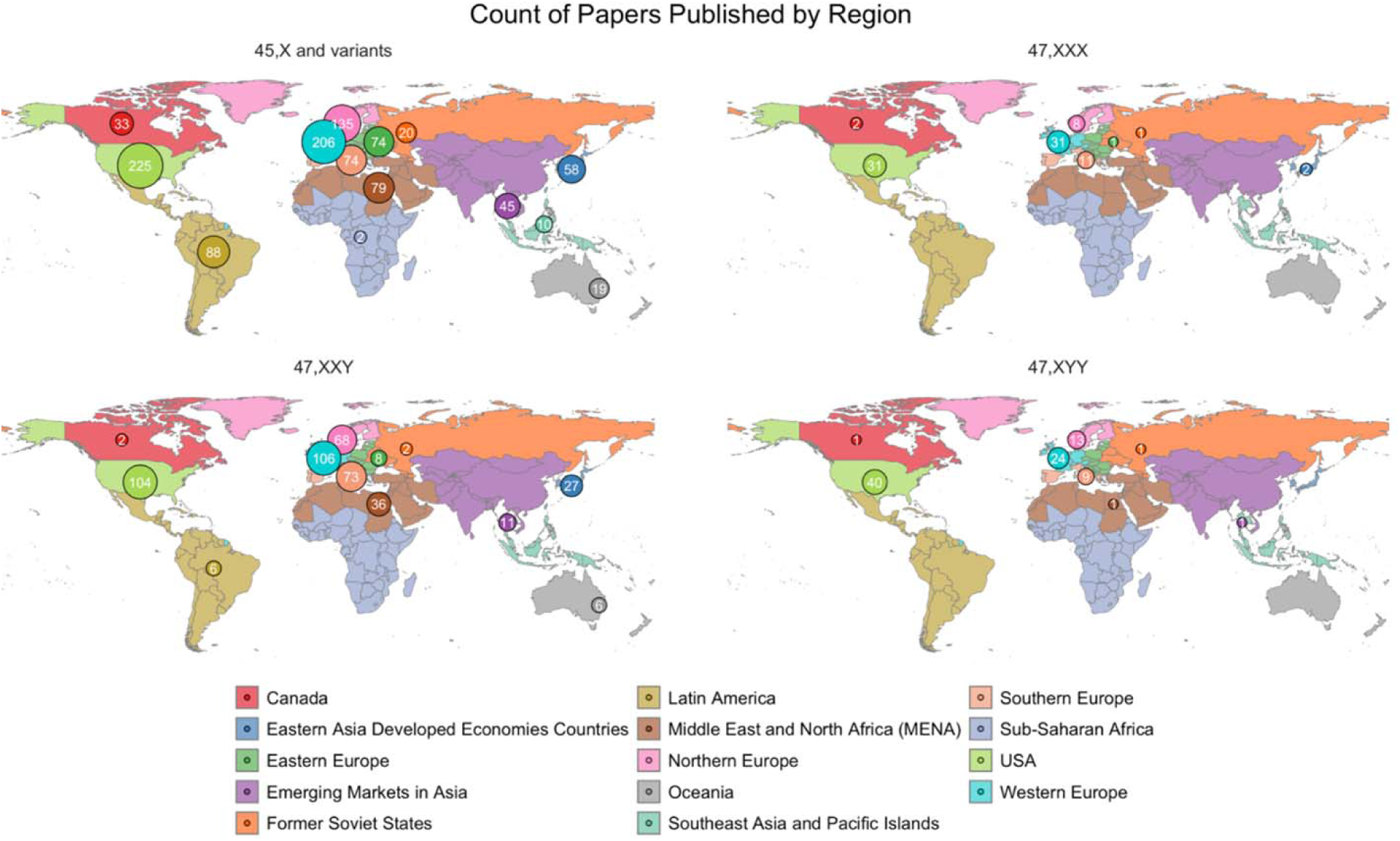
Global distribution of SCA research varies by karyotype, with prominent areas in the U.S. and Europe. Circled numbers indicate the number of manuscripts from that region.

### United States Regional Distribution

A more granular analysis of research locations within the U.S. (Figure 3) reveals regional centers for SCA research in Maryland, California, Colorado, New York, Pennsylvania, and Florida. TS research was more geographically dispersed compared to research for all other karyotypes.

**Figure 3.**
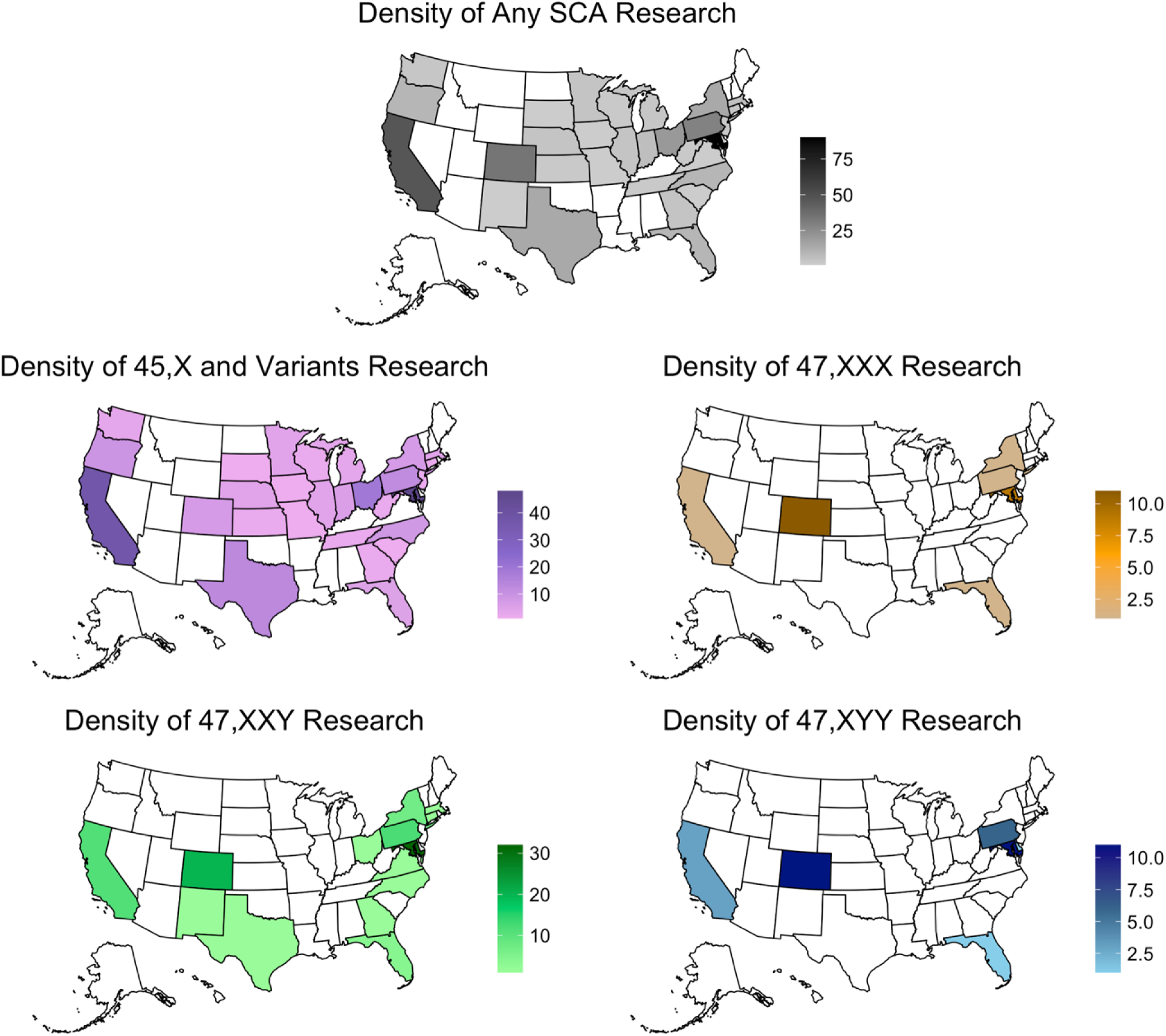
U.S.-based distribution of 329 SCA clinical research manuscripts varies by karyotypes, with prominent states including Maryland, California, Colorado, New York, Pennsylvania, and Florida. Legends indicate the number of clinical research manuscripts by state.

### Trends in Inclusion of Demographic Data

Globally, 14.3% of manuscripts reported race and/or ethnicity of study participants. This was significantly higher for U.S. papers (34.4%) compared to non-US international papers (7.9%) (p < 0.001). Reporting rates of race or ethnicity for U.S. papers increased over the 20-year observation period, with an average increase of 1.5% ± 0.4% per year (p = 0.003, Figure 4). After NIH reporting guidelines were established in 2013^13^, reporting rates of race or ethnicity for U.S. papers significantly increased from a mean of 26% ± 12.7% to 40.2% ± 14.9% per year. There was not an observed trend over time for international studies.

**Figure 4.**
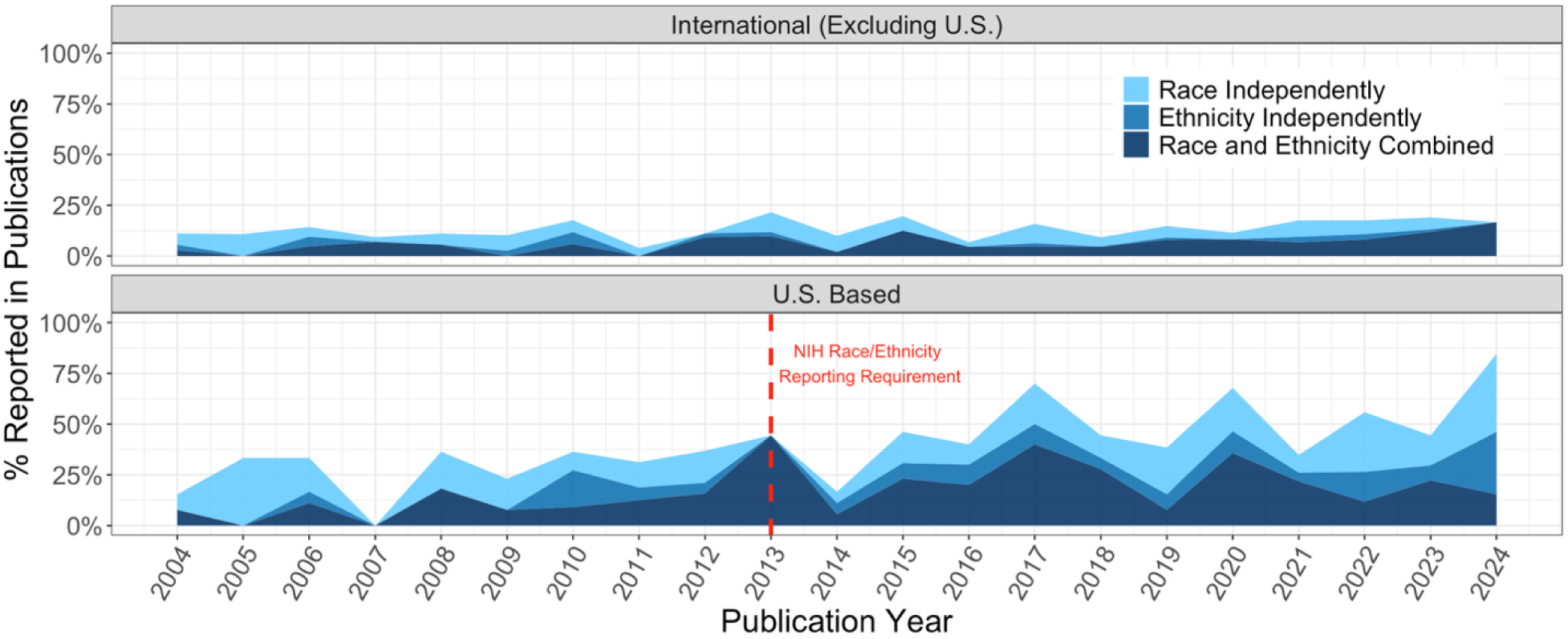
Stacked line graphs over time reveal the inclusion of race and/or ethnicity in SCA research is increasing from 2004-2024 in the U.S., though international studies are stable. The 2013 NIH guidelines on reporting race and ethnicity in research is indicated with the red dashed line.

### Comparison to U.S. Census Demographics

For the U.S. studies that did report race and/or ethnicity, the pooled proportion of White Non-Hispanic was significantly higher than that of the U.S. population^12^, while the proportion of Hispanic and Black participants were significantly lower (Table 3). The overall heterogeneity, as measured with I^2^, of proportions for race and/or ethnicity and SES across studies was generally high, ranging from 86.2% to 96.4% indicating that between-study variability was substantially higher than within-study variability (Figure 5). While, on average, our included studies differed from the U.S. estimate for White Non-Hispanic, Hispanic, and Black participants, the between-study variability is high, indicating that non-included or future studies could still be within range of the census report. When broken up by SCA, trends stay generally consistent with significantly higher proportions of White Non-Hispanic and lower proportions of Black or African American participants compared to the U.S. census (Supplemental Table 3). Significantly lower proportions of Hispanic and Asian participants were seen in 47,XXY and All Other SCT cohorts, but not in 45,X included cohorts. While I^2^ dropped as low as 3.1% for the All Other SCT cohorts, this low heterogeneity could be attributable to the smaller number of studies, putting more weight on within-study variance relative to between-study variance.

**Figure 5.**
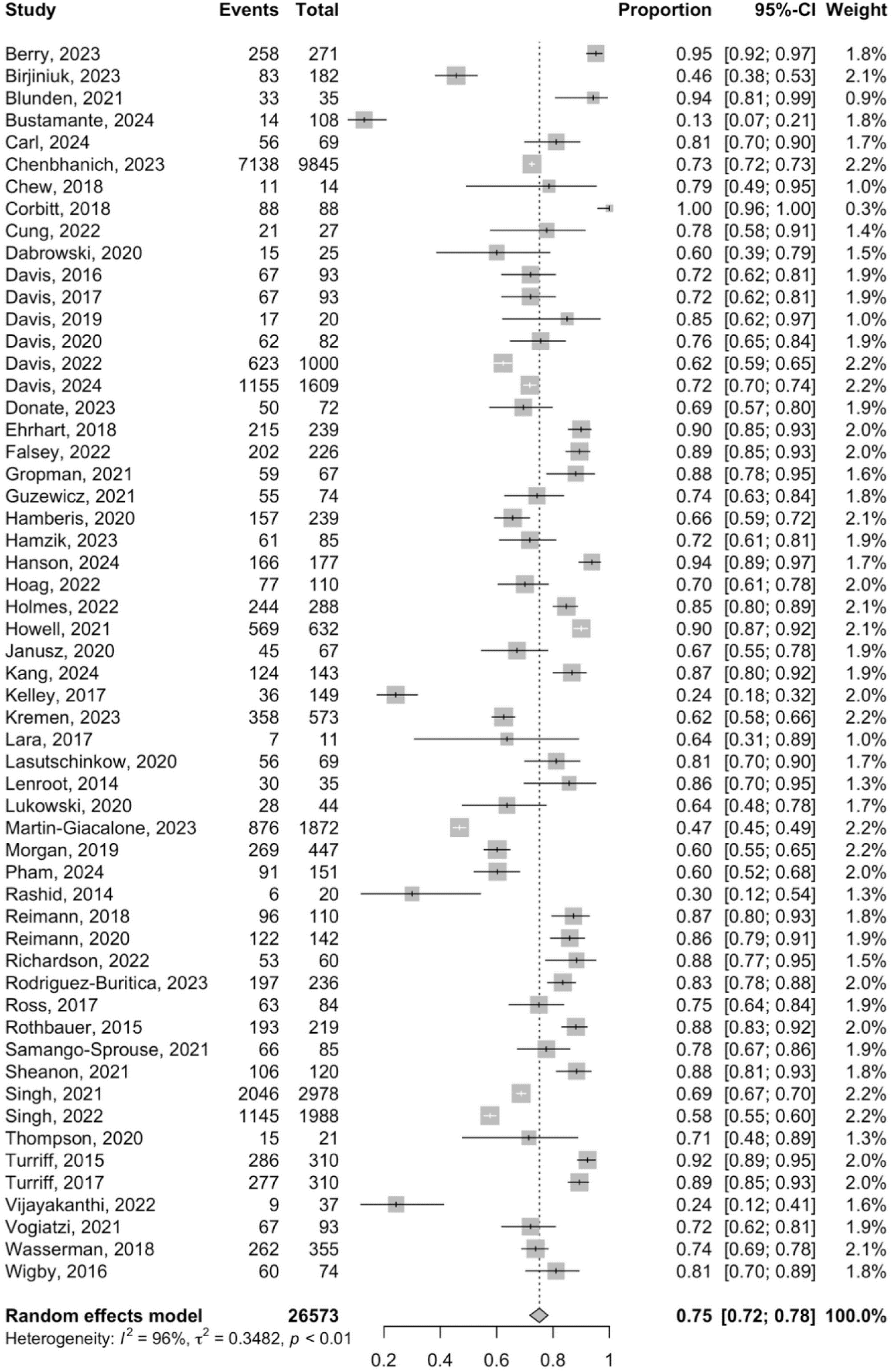
Forest plot depicting meta-analysis results for all papers, representing all SCAs, which report White Non-Hispanic proportions between 2014 and 2024. Events represent the number of White Non-Hispanic included in the study. Total represents the total number of people included in the study, factoring out participants whose race/ethnicity were reported as unknown. Weights are calculated as the inverse of the variance such that papers with higher sample sizes and therefore lower variability are given higher weights. Points and error bars represent the proportion and 95% confidence intervals, respectively, of White Non-Hispanic participants. The Random Effects Model presents the pooled proportion and 95% confidence interval, 0.75 [0.72, 0.78]. Heterogeneity measures the variability of estimates across studies, where an I^2^ estimate closer to 100% indicates substantial between-study variability relative to within-study variability.

**Table 3.** Meta-Analysis of Race/Ethnicity and SES Proportions for SCA Studies in the U.S. Compared to the 2020 U.S. Census. ^12^

| | U.S. Estimate | # of Studies | Pooled Percentage (95% CI) | $I^2$ value | p-value |
| --- | --- | --- | --- | --- | --- |
| **Race/Ethnicity** |  |  |  |  |  |
| White Non-Hispanic | 57.8% | 56 | 75.1% (71.8%, 78.2%) | 96.4% | <0.001 |
| Hispanic | 19.5% | 49 | 12.5% (10.2%, 15.3%) | 93.4% | <0.001 |
| Black or African American | 13.7% | 55 | 7.4% (6.3%, 8.6%) | 86.2% | <0.001 |
| Asian | 6.4% | 21 | 4.1% (2.6%, 6.5%) | 93.0% | 0.060 |
| **College Degree or Greater** | 37.0% | 19 | 54.6% (42.6%, 58.4%) | 92.8% | 0.002 |

### Socioeconomic Status (SES) Reporting Trends

In the last 20 years, 11.1% of total papers have presented one or more SES metric for participants, which was significantly higher for U.S. compared to non-U.S. papers (25.4% vs 8.9%, p<0.001). Figure 6 demonstrates that though there is not a statistically significant increase in reporting SES for U.S.-based papers, there is a general upward trend. Results show a <1% increase (0.65% ± 0.4%, p=0.088) in SES presentation, with a nadir in 2007 and a peak in 2015.

**Figure 6.**
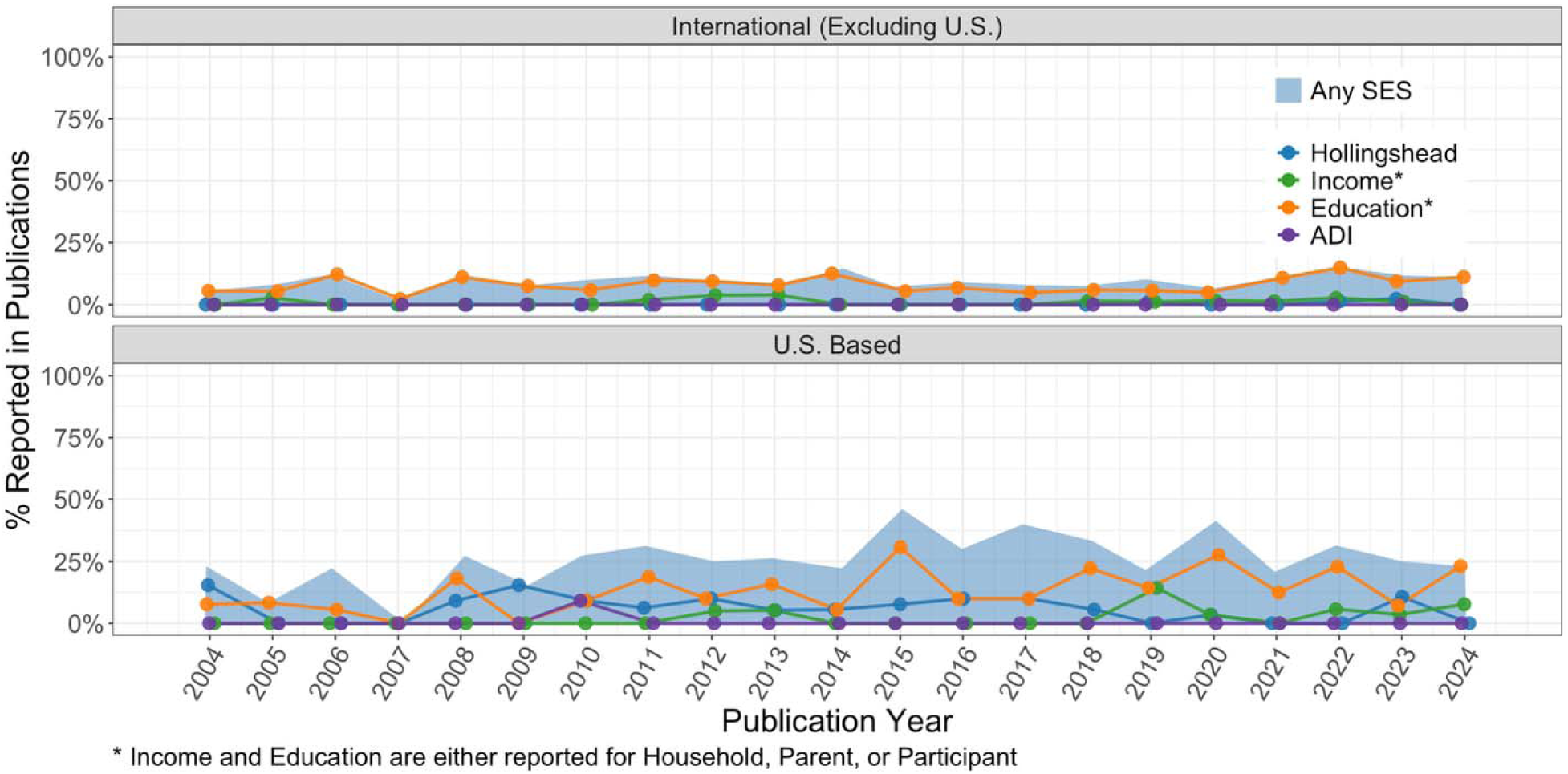
U.S.-based studies trend towards an increase in SES metrics in SCA research from 2004-2024 whil international studies remain stable.

**Figure 7.**
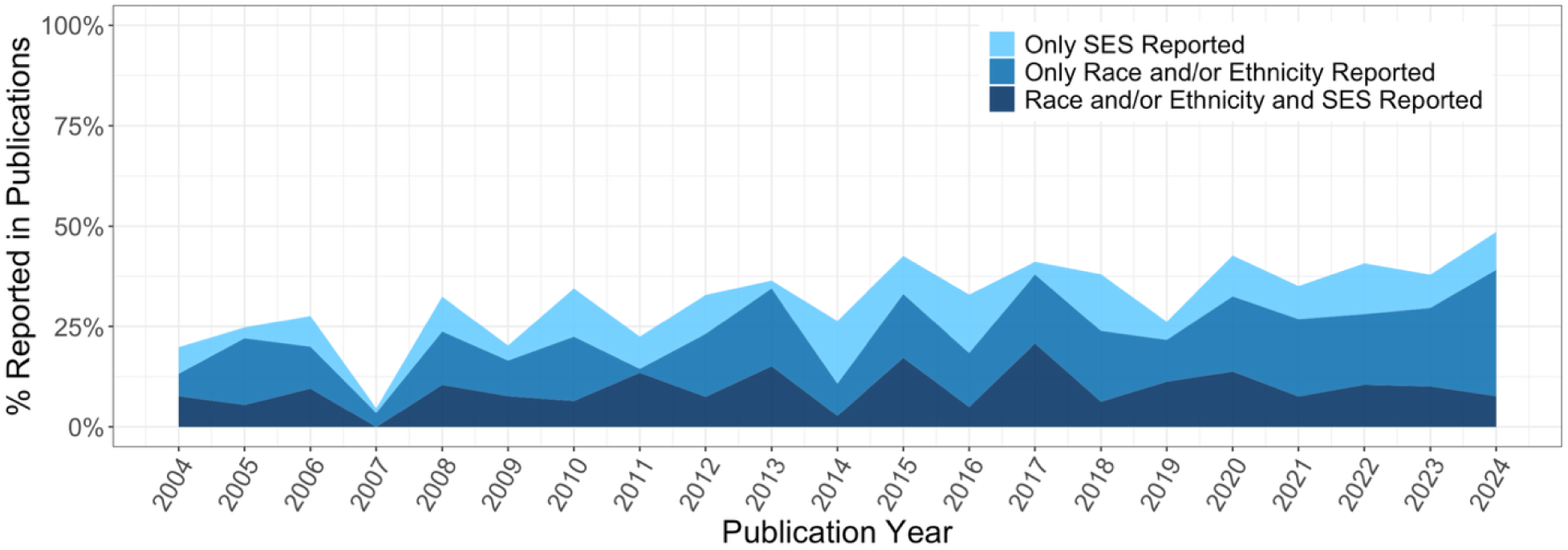
U.S.-based studies trend towards an increase in reporting of SES, and race/ethnicity, but not both, in SCA research from 2004-2024.

For studies within the U.S. reporting one or more SES metric, many used custom categorical or ordinal scales (e.g. income brackets or 0 for primary school to 3 for university education).^16^ The lack of uniform metrics limited pooled analyses across studies for statistical comparison to national benchmarks. Educational attainment (either for the participant or the parent(s) of a child participant) was the most common SES metric presented (10.5%). Of the 19 papers that presented parental educational attainment, the pooled and weighted mean percentage for college degree or higher was 54.6% (42.6%, 58.4%), which is significantly higher than the 2020 U.S. census rates^15^ of 37.0% for bachelor’s degree or higher (p=0.002). A minority of U.S.-based studies utilized standardized indices such as the Hollingshead Four-Factor Index (2.9%) and only one paper presented the Area Deprivation Index. Of the 20 papers which reported Hollingshead Index, the pooled and weighted mean (95% CI) estimate was 50.1 (46.7, 53.5), which is equivalent to the second highest social strata.

### Demographics as Analytical Variables

Of the 160 U.S. studies that presented any race/ethnicity or SES variable, 27 included the race/ethnicity or SES metrics in their analyses. Eight papers matched their control populations on race/ethnicity (3), SES (3), or both (2).^20–27^ 12 studies incorporated race/ethnicity (4), SES (4), or both (4) as covariates in analytic models but did not report results specific to these variables.^28–39^ Seven studies incorporated race/ethnicity (3), SES (3), or both (1) in their analysis and presented results about the impact of this metric on relevant outcomes (Table 4).^40–46^ Among these, 2 found no impact of the demographic metric(s) while 5 did find significant impacts.

**Table 4.**
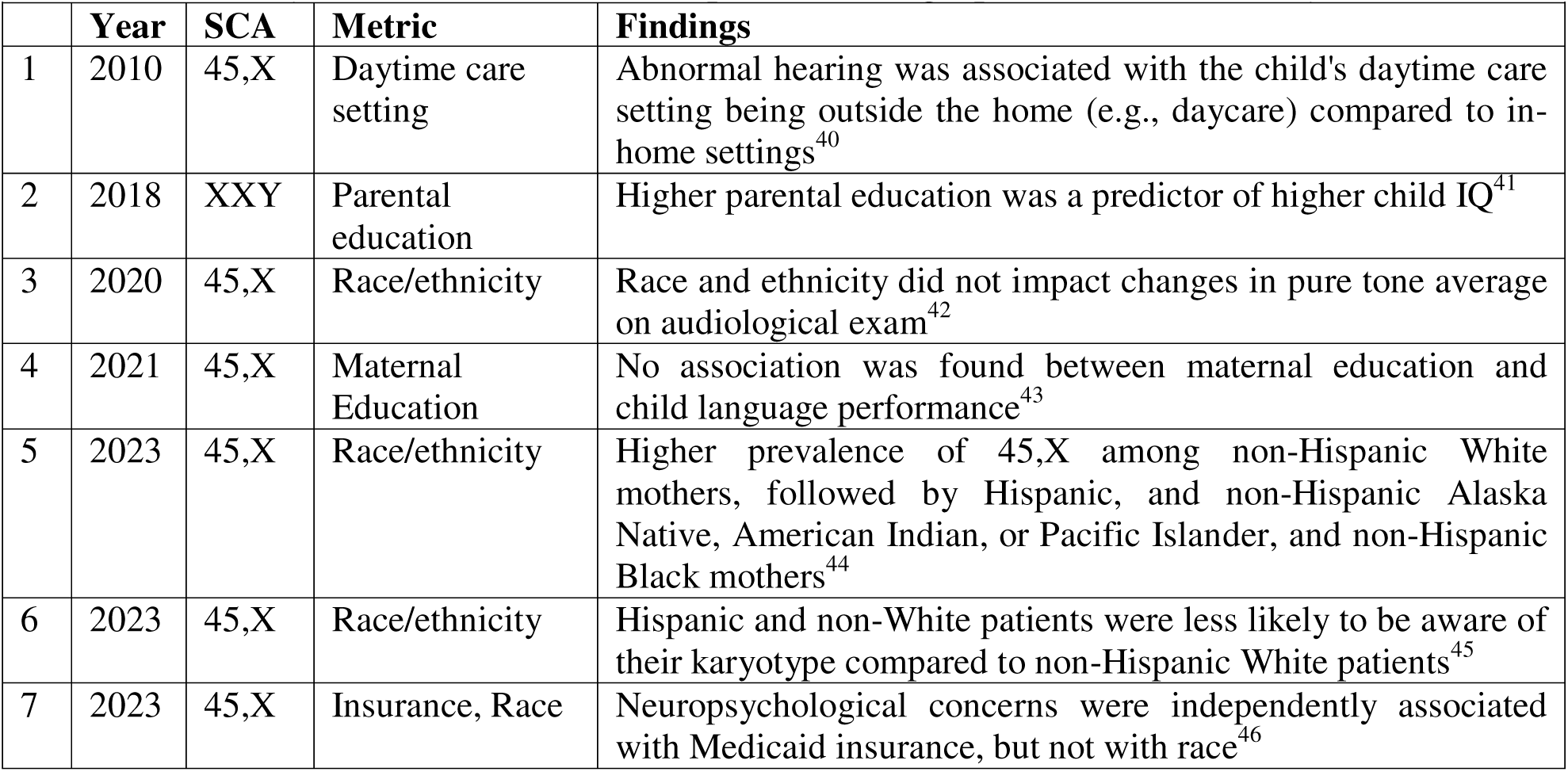
Summary of SCA studies that incorporated demographic variables in analyses.

## DISCUSSION

This systematic review provides a comprehensive overview of clinical research on SCA conditions across the world, with a focus on participant demographics and geographic representation. Our findings suggest that the existing SCA literature does not accurately reflect the population of individuals living with these conditions. Studies are heavily weighted toward TS, concentrated in the U.S. and Western Europe, and reflect an imbalanced distribution of (or mostly reflect) White, affluent, educated participants. These gaps have significant implications for the generalizability of findings, development of evidence-based guidelines, and equity in SCA research.

A major limitation of the current SCA research landscape is the inconsistent reporting of participant demographics. Despite the heterogeneous population within the U.S., fewer than half of U.S.-based studies included information on race, ethnicity, or SES. While reporting improved over time, potentially influenced by the 2013 mandate requiring the inclusion of race and ethnicity in NIH-funded studies, there remains substantial room for progress.^47^ Comprehensive reporting of demographic variables is essential for understanding how health outcomes may vary across populations and for identifying disparities in diagnosis, access to care, and treatment response.^48^ Without this information, it is impossible to determine whether clinical research is inclusive, study samples reflect the broader population, and observed findings can be generalized to all individuals with SCAs.

Among the U.S.-based studies that did report participant demographics, we observed striking underrepresentation of racially, ethnically, and socioeconomically diverse populations. Compared to U.S. census data, study populations were disproportionately composed of non-Hispanic White participants and individuals from more affluent, highly educated backgrounds. These findings are especially concerning given that SCAs arise from random chromosomal errors during meiosis and, unlike many genetic or environmentally acquired conditions, should occur with roughly equal frequency across all racial, ethnic, and socioeconomic groups.^49^ Disparities in study representation likely reflect structural barriers in access to healthcare, inequities in genetic testing, and differences in referral patterns for specialty care and research.^50^ As prenatal screening becomes more widespread, with increasing diagnoses in infancy and early childhood, it is essential that research infrastructure be prepared to engage a broader and more diverse participant base.^51^ Ensuring representative research is not only a matter of scientific rigor but a matter of health equity and justice.

Multiple factors may contribute to the demographic skew observed in SCA studies. Families with greater resources, financial, educational, or geographic, are more likely to receive a timely diagnosis, access specialized care, and engage in longitudinal research.^52,53^ Meanwhile, underdiagnosis remains a major challenge in SCAs, particularly among the trisomy conditions (47,XXY, 47,XXX, 47,XYY), where rates of identification are estimated at only 10–25%.^54^ Cultural differences in awareness and acceptance of genetic testing, language barriers, and historical mistrust of the medical system may further reduce research participation among minoritized groups.^55^ At the same time, participation in clinical research can feel overwhelming for families already navigating a new diagnosis, especially those facing additional socioeconomic or cultural stressors. Without intentional strategies to address these barriers, disparities in SCA research will persist, and may widen, as diagnosis rates increase.

The overrepresentation of 45,X in the literature also warrants mention. While 45,X is a clinically important condition with unique care needs, it has received disproportionate attention relative to the more prevalent trisomy conditions. This imbalance likely reflects a higher diagnosis rate, more established clinical care infrastructure, and research funding and priorities. Future research should aim for greater inclusion across all SCA karyotypes, with attention to the distinct developmental trajectories and healthcare challenges associated with each condition.

In sum, this systematic review highlights a critical need for more equitable and inclusive clinical research practices in the field of SCAs. As the landscape of diagnosis continues to shift, particularly with the rise of cfDNA, clinical research must evolve to capture the diversity of those affected. Improving demographic reporting, reducing barriers to participation, and expanding access to diagnosis and specialty care are essential steps toward that goal.

## LIMITATIONS

This systematic review has several limitations. First, we restricted our search to studies published in English, which may have excluded relevant non-English literature and limited the geographic diversity of the included studies. Second, our ability to evaluate demographic representation was constrained by inconsistent reporting across studies, particularly regarding SES. Many studies that reported SES used imprecise or proxy measures (e.g., education or income groupings), and only a minority included validated instruments. Third, our analysis was based on unique publications rather than unique cohorts; as a result, some studies may be overrepresented due to multiple papers derived from the same participant sample. Without standardized identifiers for study populations, we were unable to adjust for this potential duplication. In addition, while we aimed to capture original clinical research, our inclusion criteria excluded small case reports and conference abstracts, which may have included data from underrepresented populations. Finally, we were limited to evaluating reported data; we could not assess participant demographics in studies that did not report them, potentially over or underestimating the extent of underrepresentation.

## CONCLUSIONS

This systematic review reveals gaps in the demographic reporting and representation of participants in SCA research, with overrepresentation of non-Hispanic White and higher SES individuals. The lack of demographic data across much of the literature limits the ability to assess generalizability, and applicability of findings to the broader population of individuals with SCAs. As research advances and diagnostic practices evolve, there is a critical need to ensure demographic and socioeconomic variation that reflects the population affected by SCA is prioritized, reported, and analyzed. Future research should adopt standardized demographic reporting practices, develop intentional recruitment strategies for underrepresented populations, and build infrastructure to support equitable access to clinical research. Closing these gaps is essential to ensuring that research translates into inclusive, evidence-based care for the full spectrum of individuals with SCAs.

## Data Availability

Data will be made available upon reasonable request to the authors.

---

**Supplemental Table 1.**
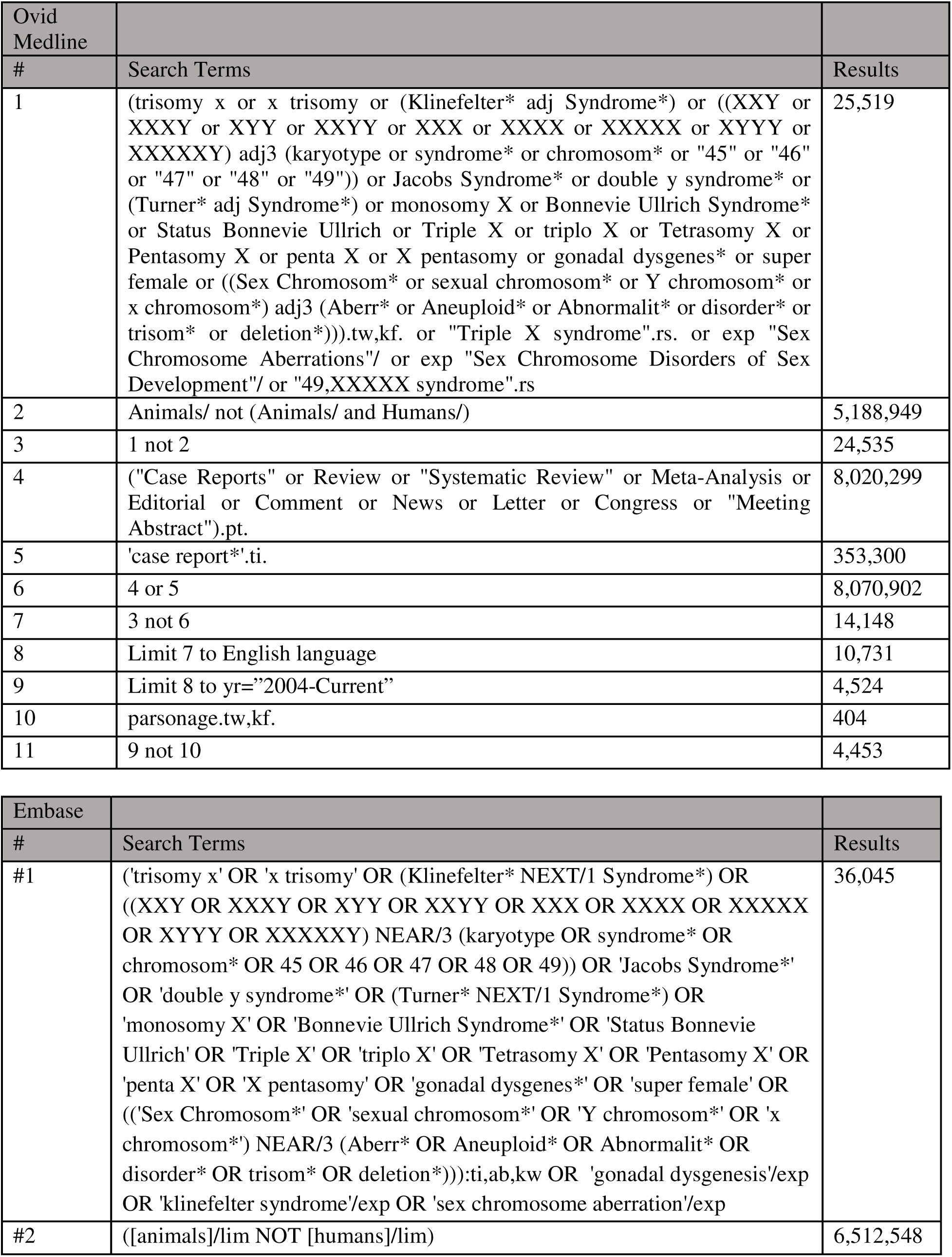
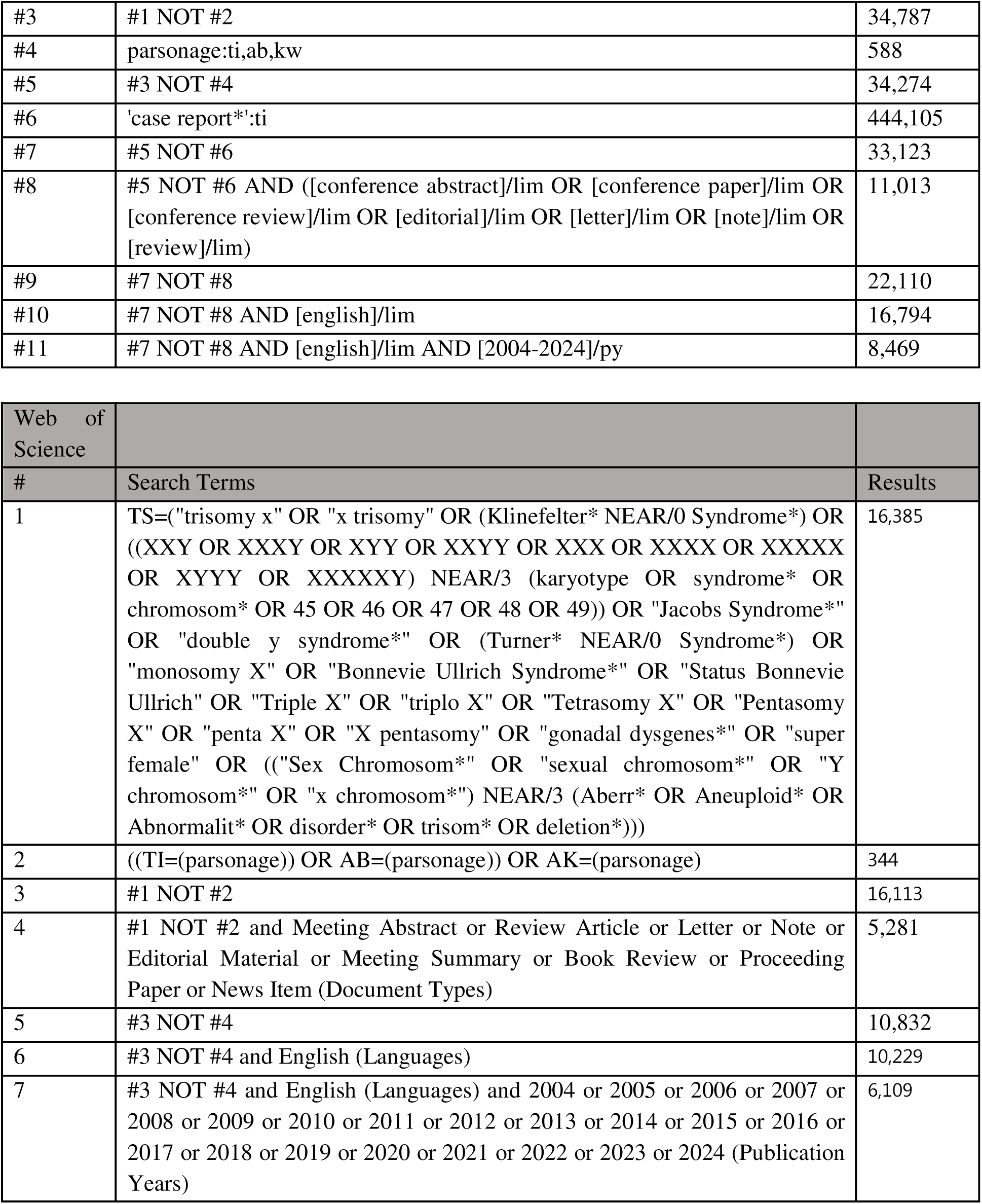

**Supplemental Table 2.** Topic of research by karyotype.

| **Topic of study** | **45,X**<br>**(N = 994)** | **47,XXY**<br>**(N = 426)** | **47,XYY**<br>**(N = 78)** | **47,XXX**<br>**(N = 75)** | **48,XXYY**<br>**(N = 28)** | **Other**<br>**(N = 21)** |
| --- | --- | --- | --- | --- | --- | --- |
| Cardiac /<br>Cardiometabolic | 203 (20.4%) | 32 (7.5%) | 2 (2.6%) | 2 (2.7%) | 1 (3.6%) | 1 (4.8%) |
| Neurodevelopment /<br>psych / mental health | 89 (9%) | 113 (26.5%) | 48 (61.5%) | 45 (60%) | 14 (50%) | 9 (42.9%) |
| Physical phenotype | 151 (15.2%) | 45 (10.6%) | 8 (10.3%) | 8 (10.7%) | 4 (14.3%) | 2 (9.5%) |
| Other or multiple | 139 (14%) | 48 (11.3%) | 9 (11.5%) | 8 (10.7%) | 5 (17.9%) | 4 (19%) |
| Growth treatment | 149 (15%) | 0 (0%) | 0 (0%) | 0 (0%) | 0 (0%) | 0 (0%) |
| Fertility / fertility<br>treatment | 43 (4.3%) | 96 (22.5%) | 4 (5.1%) | 4 (5.3%) | 1 (3.6%) | 2 (9.5%) |
| Genetics / epigenetics | 89 (9%) | 28 (6.6%) | 3 (3.8%) | 3 (4%) | 2 (7.1%) | 0 (0%) |
| Sex steroid hormones /<br>hormone treatment | 61 (6.1%) | 39 (9.2%) | 2 (2.6%) | 0 (0%) | 0 (0%) | 1 (4.8%) |
| Quality of Life /<br>Wellbeing | 65 (6.5%) | 17 (4%) | 2 (2.6%) | 4 (5.3%) | 1 (3.6%) | 1 (4.8%) |
| Basic science | 5 (0.5%) | 8 (1.9%) | 0 (0%) | 1 (1.3%) | 0 (0%) | 1 (4.8%) |

**Supplemental Table 3.** Meta-Analysis of Race/Ethnicity and SES Proportions by SCA Compared to the 2020 U.S. Census.

|  | US<br>Estimate<sup>10</sup> | 45,X |  |  | 47,XXY |  |  | All Others |  |  |
| --- | --- | --- | --- | --- | --- | --- | --- | --- | --- | --- |
|  |  | # | I<sup>2</sup> | Pooled<br>Percentage<br>(95% CI) | # | I<sup>2</sup> | Pooled<br>Percentage<br>(95% CI) | # | I<sup>2</sup> | Pooled<br>Percentage<br>(95% CI) |
| Race/Ethnicity |  |  |  |  |  |  |  |  |  |  |
| White Non-Hispanic | 57.8% | 32 | 97.0% | 68.3%***<br>(63.3%, 72.8%) | 15 | 92.7% | 78.3%***<br>(72.1%, 83.5%) | 10 | 64.7% | 87.0%***<br>(82.5%, 90.5%) |
| Hispanic | 19.5% | 28 | 93.4% | 17.6%<br>(14.0%, 21.8%) | 12 | 72.6% | 8.9%***<br>(6.7%, 11.7%) | 8 | 3.1% | 5.6%***<br>(3.9%, 8.1%) |
| Black or African American | 13.7% | 34 | 84.8% | 7.3%***<br>(6.0%, 9.0%) | 14 | 83.9% | 8.7%***<br>(6.6%, 11.3%) | 7 | 54.8% | 4.4%***<br>(2.3%, 8.3%) |
| Asian | 6.4% | 20 | 93.3% | 4.2%<br>(2.6%, 6.7%) | 11 | 63.0% | 2.9%***<br>(2.1%, 4.0%) | 6 | 40.4% | 3.3%*<br>(1.7%, 6.4%) |
| College Degree or Higher | 37.0% | 9 | 85.1% | 47.2%**<br>(39.1%, 55.4%) | 6 | 96.3% | 51.0%<br>(31.0%, 70.6%) | 4 | 80.1% | 59.8%*<br>(39.6%, 77.1%) |

\*\*\*: $p < 0.001$ , \*\*: $p < 0.01$ , \*: $p < 0.05$ . p-values represent one-sample z-tests compared to the US census report.

